# Effectiveness of a pharmacy-led personalised intervention program aimed at improving adherence to oral diabetes and/or hypertension medication in people with type 2 diabetes

**DOI:** 10.64898/2026.10.28.26366703

**Authors:** Marlous Langendoen-Gort, Sharon Remmelzwaal, Femke Rutters, Hiyam Al-Jabr, Maartje de Wit, Debi Bhattacharya, Jacqueline G Hugtenburg, Andrew Farmer, Petra JM Elders

## Abstract

**Aims/hypothesis:** To evaluate the effectiveness of a personalised intervention program aimed at improving medication adherence in people with type 2 diabetes who are non-adherent to oral diabetes and/or hypertension medication, compared to usual care including access to a diabetes website, after a six-month follow-up.

**Methods:** Parallel-group randomised controlled trial with six-month follow-up conducted in community pharmacies in the Netherlands and United Kingdom. People with type 2 diabetes, 35-75 years and non-adherent to oral diabetes and/or hypertension medication were recruited. The intervention group received the personalised intervention program and the control group received usual care and an informative diabetes website. The primary outcome was medication adherence measured by telephone pill count and secondary outcomes were self-reported medication adherence, systolic blood pressure and HbA1c level. Linear regression analysis and longitudinal mixed model analysis were performed to assess intervention effectiveness.

**Results:** The study population comprised 68 participants, 66.2% men with a median age of 65.0 [Q1 to Q3 59.3 to 71.0] years. Linear regression analysis for the primary outcome medication adherence showed no significant effect of the intervention (B −0.014, 95% CI −0.17 to 0.14). In addition, no significant differences were found between the intervention and control group for self-reported medication adherence (OR 3.18, 95% CI 0.89 to 11.37) and systolic blood pressure (B −1.41, 95% CI −9.64 to 6.83). Finally, the intervention group did not show better HbA1c levels compared to the control group (B 0.68, 95% CI 0.44 to 0.92).

**Conclusions/interpretation:** We did not observe a statistically significant difference, nor a clinically point estimate of difference, in medication adherence between the intervention and control group of a personalised intervention program aimed at improving medication adherence in people with type 2 diabetes who are non-adherent to oral diabetes and/or hypertension medication after six-months follow-up. We therefore do not recommend implementation of the intervention program in daily practice.

**Trial registry number:** Registered in the Dutch Trial Register (Trial NL8747) and ISRCTN Registry (ISRCTN36009809).

**Social media post:** No effect of a personalised intervention program was observed on medication adherence in people with type 2 diabetes. Future studies should first focus on alternative recruitment methods for non-adherent people with type 2 diabetes, before the effectiveness of the intervention can be re-evaluated in a larger study sample.

**Research in context:** *What is already known about this subject?:*

- Medication non-adherence is a common health problem in people with type 2 diabetes.
- Most medication adherence interventions showed small and inconsistent effects, potentially because interventions are not personalised to the needs of participants.

*What is the key question?:*

- What is the effect of a personalised intervention program aimed at improving medication adherence in people with type 2 diabetes who are non-adherent to oral diabetes and/or hypertension medication, compared to usual care including access to a publicly available diabetes website, after six months of follow-up?

*What are the new findings?:*

- No effect was observed of the personalised intervention program on medication adherence measured with a telephone pill count.
- No effect was observed of the personalised intervention program on self-reported medication adherence and systolic blood pressure.
- The intervention group did not show better HbA1c levels compared to the control group.
- Recruitment of participants was challenging and as such we were unable to achieve the intended study sample.

*How might this impact on clinical practice in the foreseeable future?:*

- We currently do not recommend implementation of the personalised intervention program in routine practice. Further work is first needed to establish alternative recruitment methods for non-adherent people with type 2 diabetes, before the effectiveness of the intervention can be re-evaluated in a larger study sample.

## Introduction

Medication non-adherence is a common health problem in people with type 2 diabetes (1). To date, many interventions aimed at improving medication adherence have been developed, but these often showed small and inconsistent effects (2). A potential cause of these limited effects is that interventions are not tailored to the barriers for medication adherence (2, 3).

Therefore, in this study we evaluate the effect of a personalised intervention program that is aimed at improving medication adherence in people with type 2 diabetes who are non-adherent to oral diabetes and/or hypertension medication, compared to usual care including access to a publicly available diabetes website, after six-months of follow-up.

## Methods

Approval by the medical ethical committee of Amsterdam UMC location VUmc and HRA and Health and Care Research Wales was obtained. Written informed consent was collected (4).

### Study design

A parallel-group randomised controlled trial, with a six-month follow-up, was performed in pharmacies in the Netherland and United Kingdom (UK). Participants were included between October 2020 and February 2022 and were randomised to the intervention or control group. Details of the study design can be found in the study protocol (4).

### Study population and setting

People with type 2 diabetes, non-adherent to oral diabetes and/or hypertension medication, 35-75 years, mobile phone user and able to understand text messages were recruited. Exclusion criteria were invalid medication adherence data, use of pharmacies’ pill packaging service and major psychiatric disorder illness. In the Netherlands, people who started or stopped using diabetes medication during the period in which the dispensing score was calculated were also excluded. In the UK, people using insulin were excluded.

Non-adherent people were identified with an in-practice electronic search. This was conducted in dispensing records of pharmacies (the Netherlands) or prescribing records of general practices (UK). A dispensing score < 90% over a one-year period and a prescribing score < 80% over a two-year period were considered non-adherent.

### Intervention

#### Intervention group

The intervention group received the personalised intervention program. Participants completed a questionnaire to identify barriers for medication adherence. Based on these barriers participants were allocated to non-adherence profile(s), corresponding to a pre-defined set of supporting modules. Available supporting modules were brief messages, clinical medication review, medication schedule, reminders, medication dispensing systems, smart messages, referral to general practitioner, and web-based self-help application for low mood (**Additional files, Table A1**). Thereafter, participants had an appointment with their pharmacist for shared-decision making on the final set of supporting modules.

#### Control group

The control group received usual care and access to a publicly available diabetes website. Participants in the control group had an appointment with the pharmacy assistant in which participants were informed about the diabetes website.

### Outcome measures

#### Primary outcome

The primary outcome was medication adherence with diabetes or hypertension medication assessed with a telephone pill count (5). Medication adherence was calculated using an adapted version of a previously reported equation (6). The following equation was used: ((number of dosage units in stock at baseline pill count + number of dosage units dispensed between baseline and follow-up pill count) – number of dosage units remaining at follow-up pill count) / (prescribed number of dosage units per day × number of days between baseline and follow-up pill count). Higher scores indicate better adherence. Only medicines counted at baseline and follow-up pill counts, and with available dispensing data, were evaluated.

#### Secondary outcomes

Secondary outcomes were self-reported medication adherence, systolic blood pressure (SBP) and HbA1c level. The Medication Adherence Report Scale (MARS-5) questionnaire (7) was used to assess self-reported medication adherence. The questionnaire comprises five items scored on a five-point scale. A sum score was calculated (range 5-25) and analysed as a dichotomous sum score (i.e. 0-24 and 25) (8). To obtain participants’ SBP (mmHg) and HbA1c level (mmol/mol) registry data were consulted. In the Netherlands, SBP and HbA1c measurements were collected using prespecified time windows (4) and in the UK the latest available measurements were used at baseline and 6-month follow-up. For the UK follow-up analysis, participants were only included if a subsequent measurement was available within the 6-month timeframe.

### Statistical analysis

For baseline characteristics descriptive statistics were used. Continuous variables are presented using means and standard deviations and categorical variables using frequencies and percentages. For not normally distributed continuous variables medians and interquartile ranges are presented.

All effect analyses were performed according to an intention-to-treat principle. For medication adherence, SBP and HbA1c level, linear regression analyses were conducted. Missing SBP and HbA1c level data were imputed using multiple imputation and analyses were corrected for the baseline value of the outcome. For the MARS-5 logistic longitudinal mixed model analysis was performed. For all effect analyses, a crude analysis and an analysis assessing potential effect modifiers (i.e. age, sex and educational level) were performed. A p-value of <0.05 was considered statistically significant. SPSS Statistics version 28 was used for analyses.

## Results

### Baseline characteristics

In total, 25 community pharmacies included 68 participants. Both the intervention and control group comprised 34 participants. The overall study population consisted of 66.2% men, with a median age of 65.0 [Q1 to Q3 59.3 to 71.0] years. **Table 1** presents the baseline characteristics of the study population.

**Table 1.** Baseline characteristics of the study population.

|  | **Study population**<br>**(n=68)**<br>*N (%) or mean ± SD or*<br>*median [Q1 to Q3]* | **Intervention group (n=34)**<br>*N (%) or mean ± SD or median*<br>*[Q1 to Q3]* | **Control group (n=34)**<br>*N (%) or mean ± SD or median*<br>*[Q1 to Q3]* |
| --- | --- | --- | --- |
| **Age in years** | 65.0 [59.3 to 71.0] | 66.5 [59.8 to 73.0] | 63.0 [59.0 to 70.0] |
| **Sex** |  |  |  |
| Male | 45 (66.2%) | 26 (76.5%) | 19 (55.9%) |
| **Country of birth<sup>°</sup>** |  |  |  |
| The Netherlands | 42 (61.8%) | 21 (61.8%) | 21 (61.8%) |
| Surinam | 3 (4.4%) | 2 (5.9%) | 1 (2.9%) |
| Other | 2 (2.9%) | 1 (2.9%) | 1 (2.9%) |
| **Ethnicity<sup>–</sup>** |  |  |  |
| White British | 19 (27.9%) | 9 (26.5%) | 10 (29.4%) |
| Black or black British | 1 (1.5%) | 1 (2.9%) | 0 (0.0%) |
| Other white background | 1 (1.5%) | 0 (0.0%) | 1 (2.9%) |
| **Educational level** |  |  |  |
| High | 38 (55.9%) | 19 (55.9%) | 19 (55.9%) |

<sup>°</sup> Only reported for the Netherlands; <sup>–</sup> Only reported for the United Kingdom

### Primary outcome

**Table 2** presents the proportions and mean scores (±SD) of medication adherence for both treatment groups. The median value of medication adherence after six months follow-up was 0.96 [Q1 to Q3 0.62 to 1.03] for the intervention group and 0.88 [Q1 to Q3 0.70 to 1.01] for the control group. Linear regression analysis showed an effect of the intervention of −0.014 (95% CI −0.17 to 0.14) (**Table 3**). Effect modification by age, sex or educational level was not found.

**Table 2.** Mean scores (± SD), median scores [Q1 to Q3] or frequencies (%) of the primary and secondary outcomes according to intention-to-treat analyses.

| Outcomes |  | Baseline |  | 3-months follow-up |  | 6-month follow-up |  |
| --- | --- | --- | --- | --- | --- | --- | --- |
|  |  | N | N (%) or mean ± SD or median [Q1 to Q3] | N | N (%) or mean ± SD or median [Q1 to Q3] | N | N (%) or mean ± SD or median [Q1 to Q3] |
| Primary outcome |  |  |  |  |  |  |  |
| Medication adherence | Intervention |  |  |  |  | 24 | 0.96 [0.62 to 1.03] |
|  | Control | - | - | - | - | 29 | 0.88 [0.70 to 1.01] |
| Secondary outcomes |  |  |  |  |  |  |  |
| MARS-5 (≥25) | Intervention | 34 | 14.0 (41.2%) | 26 | 12.0 (46.2%) | 27 | 11.0 (40.7%) |
|  | Control | 33 | 7.0 (21.2%) | 28 | 8.0 (28.6%) | 29 | 6.0 (20.7%) |
| SBP (mmHg) | Intervention | 26 | 132.3 (± 12.2) | - | - | 18 | 133.3 (± 9.2) |
|  | Control | 26 | 135.5 (±14.8) |  |  | 22 | 137.4 (± 16.4) |
| HbA1c level (mmol/mol) | Intervention | 27 | 57.4 (± 9.2) | - | - | 24 | 60.4 (± 12.4) |
|  | Control | 28 | 58.6 (± 13.7) |  |  | 25 | 56.4 (± 12.2) |

Medication Adherence Report Scale (MARS-5); Systolic blood pressure (SBP)

**Table 3.**
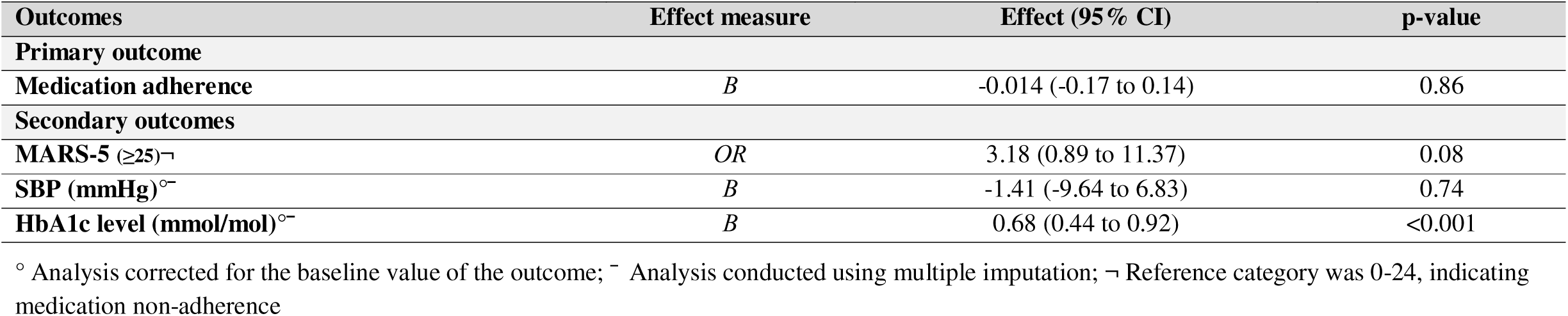
Differences between the intervention groups on the primary and secondary outcomes according to intention-to-treat analyses.

### Secondary outcomes

For the secondary outcomes, proportions and mean (±SD) or median scores [Q1 to Q3] are also presented in **Table 2**. No significant effects between treatment groups were found for self-reported medication adherence (OR 3.18, 95% CI 0.89 to 11.37) and SBP (B −1.41, 95% CI −9.64 to 6.83) (**Table 3**). Finally, participants in the intervention group did not show better HbA1c levels (B 0.68, 95% CI 0.44 to 0.92) compared to the control group (**Table 3**). Effect modification by age, sex or educational level was not found.

## Discussion

We evaluated the effect of a personalised intervention program aimed at improving medication adherence to oral diabetes and/or hypertension medication in people with type 2 diabetes. No statistically significant, nor a clinically point estimate of difference, was observed between the treatment groups for medication adherence self-reported medication adherence and SBP. In addition, participants in the intervention group did not show better HbA1c levels.

Our study was based on the CATI study, aimed at improving medication adherence in people with hypertension (9). In line with our results, CATI researchers found no statistically significant differences between treatment groups on self-reported medication adherence and blood pressure (8). Limited eligibility of the CATI participants was identified as a potential cause for the limited effect of the intervention in a process evaluation (8, 10). Therefore, we adjusted the inclusion and exclusion criteria in our study. However, we were unable to include the intended number of 300 participants.

Additionally, compared to the CATI study we used digital supporting modules. A recent meta-analysis showed that mobile phone reminders, in addition to usual care, had a positive effect on medication adherence (11). While we were unable to support this effect, participants often expressed satisfaction with the reminders (12).

### Limitations and strengths

We were unable to include the intended number of participants, restricting us in conducting multilevel analysis. Inclusion criteria were expanded during the study by raising the dispensing score from 80% to 90%. Consequently, the study population exhibited higher levels of baseline medication adherence. Missing data because participants declined measurements, technical issues and incomplete/non-standardised data collection further compounded challenges with power. Finally, ethical approval constraints prohibited data collection of potential participants who did not consent, thus we are unable to provide commentary regarding selection bias.

This study also has several strengths. First, we were able to conduct this study in both the Netherlands and the UK, even though these countries both have different health care systems, increasing the generalizability of the results. Second, we have included a large sample of pharmacies in the Netherlands, indicating pharmacy support for a pharmacy-led medication adherence intervention. Third, we were able to develop and evaluate the effect of a personalised intervention program, which based on literature seems to be more effective in improving medication adherence (2, 3). This study therefore adds on the information available about such interventions.

### Implications for future research and practice

We were unable to include the intended number of participants, hampering us in conducting the effect analysis. We think that future studies should focus on alternative methods to recruit non-adherent people with type 2 diabetes. The effectiveness of the personalised intervention program should be re-evaluated in a larger sample (12).

## Conclusion

In conclusion, in our study population we did not observe a statistically significant difference in medication adherence, self-reported medication adherence and SBP between the intervention and control group after six-months follow-up. In addition, the intervention group did not show better HbA1c levels. Therefore, we currently do not recommend implementing the personalised intervention program in daily practice.

## Data Availability

All data produced in the present study are available upon reasonable request to the authors via https://hoornstudies.com

## Acknowledgements

The authors would like to thank students that provided data-analyses support.

## Data availability

Participant data cannot be supplied because no informed consent was obtained for data sharing.

## Authors’ relationships and activities

The authors declare no conflicts of interest.

## Contribution statement

AF, JH and PE developed the basis for the study design and protocol. DB, FR and MW were involved in the study design. AF, DB, HA, JH, MLG and PE were members of the trial managing committee. MLG coordinated data collection in the Netherlands and DB coordinated data collection in the UK. MLG and SR conducted data analyses. MLG drafted this manuscript. All authors read and approved this final short communication manuscript.

## Additional files

**Table A1.**
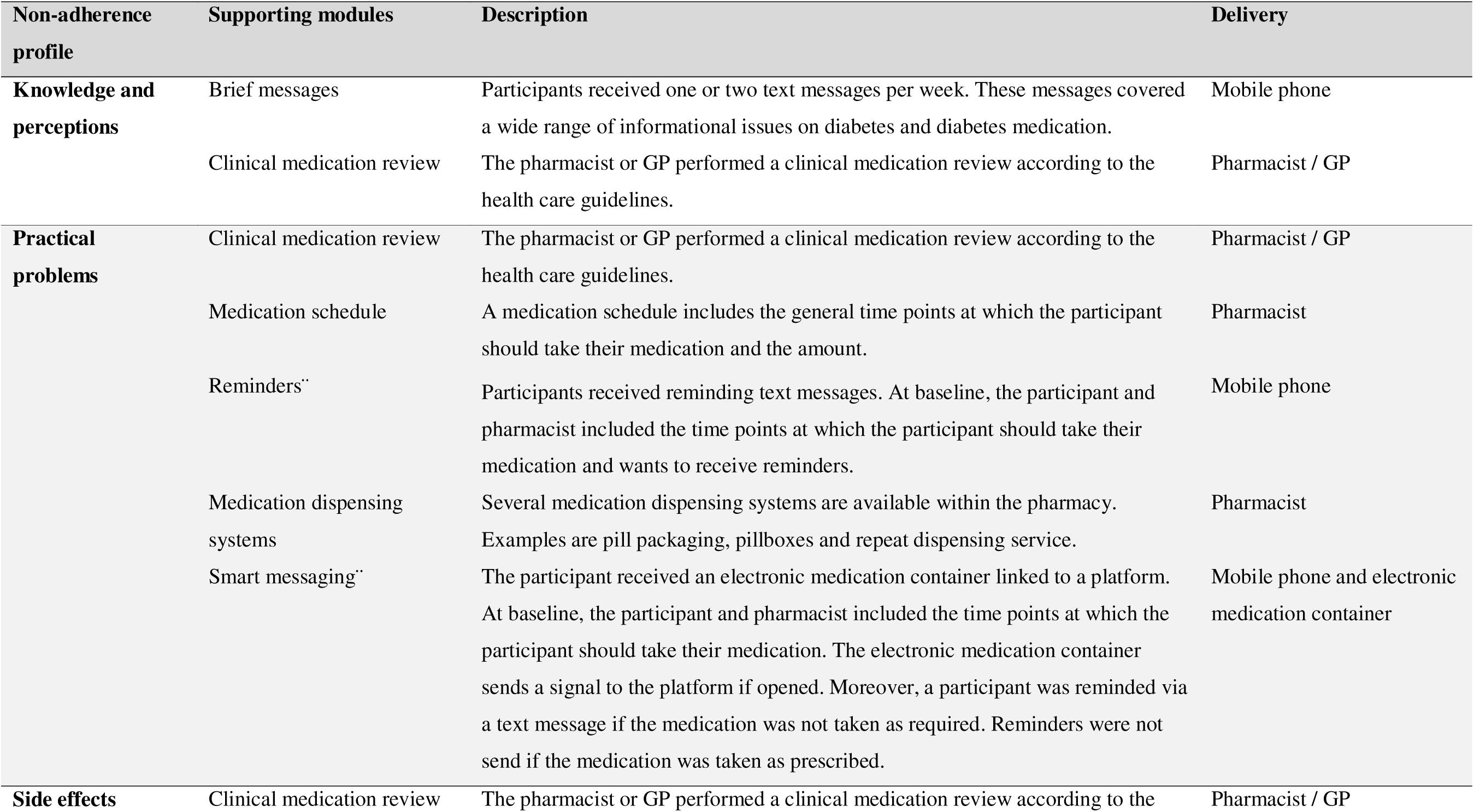
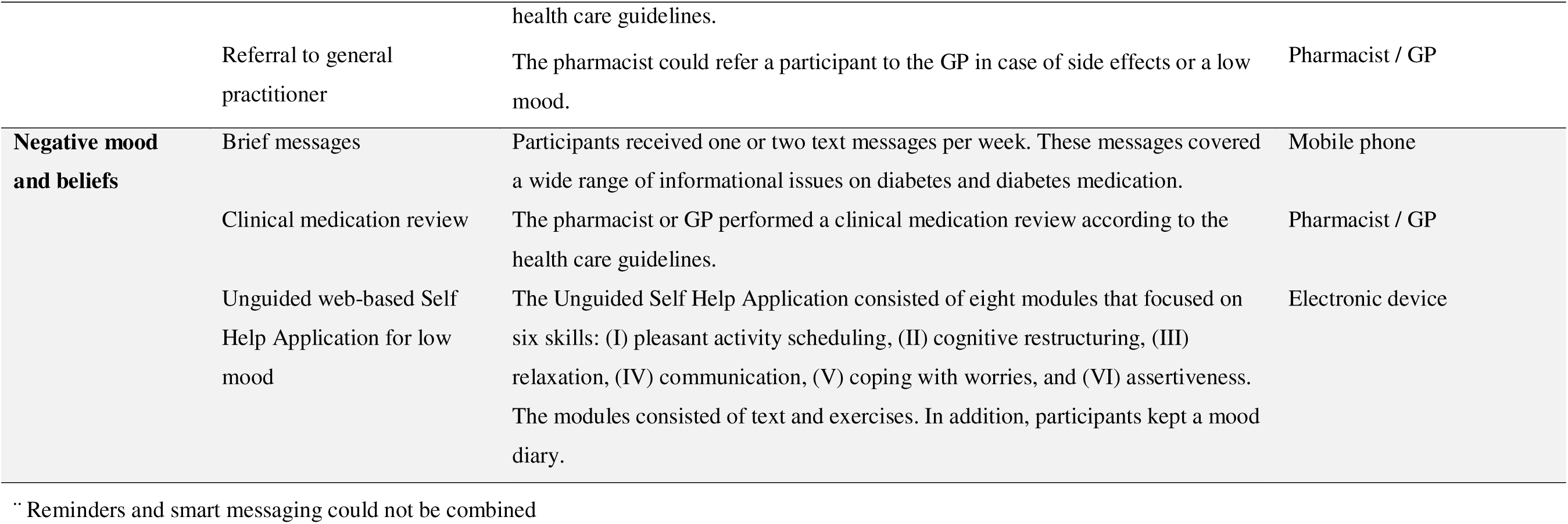
Supporting modules based on the non-adherence profiles.

**Figure A1.**
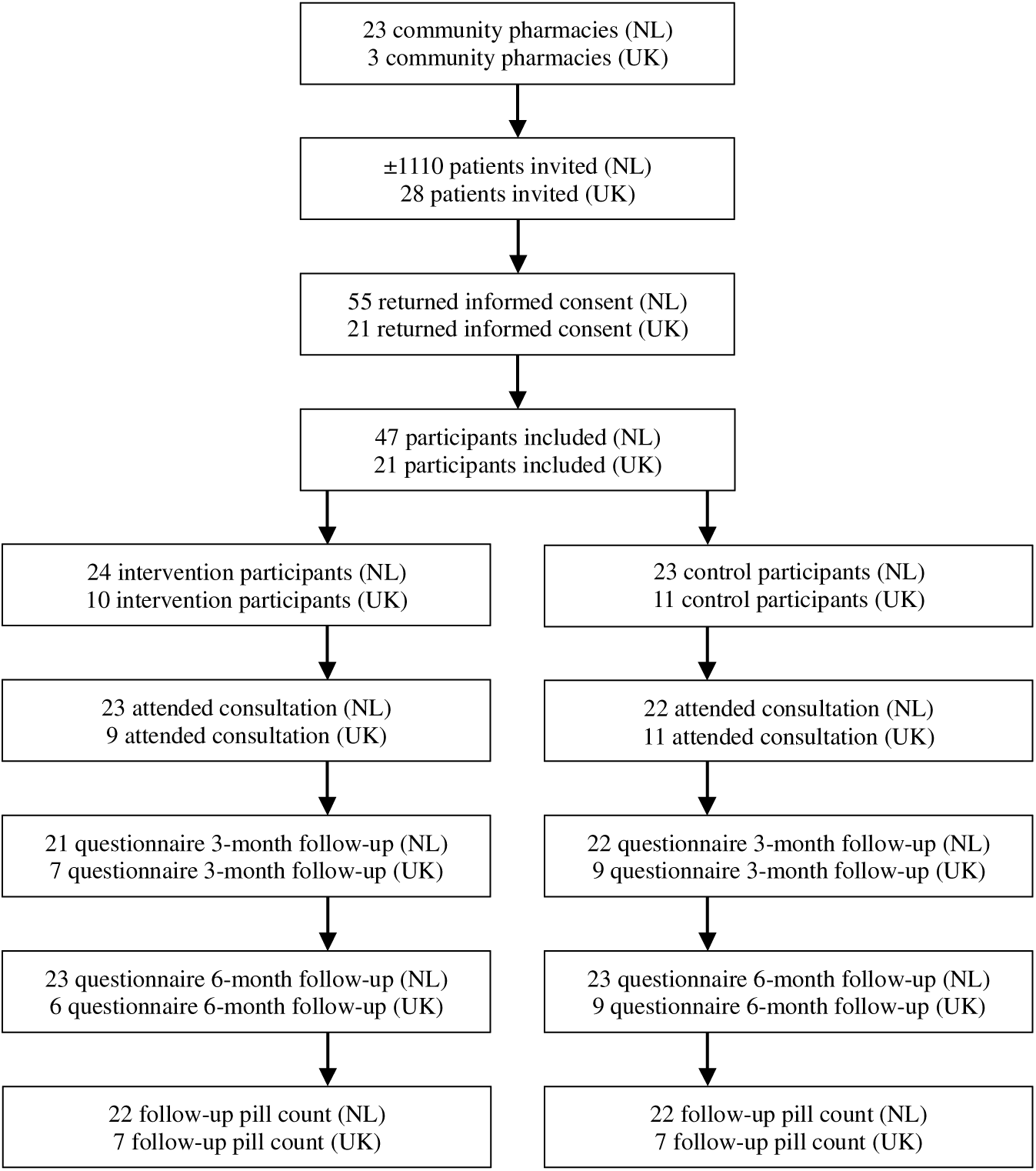
Participant flowchart

## References

1. Evans M, Engberg S, Faurby M, Fernandes J, Hudson P, Polonsky W. Adherence to and persistence with antidiabetic medications and associations with clinical and economic outcomes in people with type 2 diabetes mellitus: A systematic literature review. Diabetes Obes Metab. 2022;24(3):377–90.

2. Sapkota S, Brien JA, Greenfield JR, Aslani P. A Systematic Review of Interventions Addressing Adherence to Anti-Diabetic Medications in Patients with Type 2 Diabetes-Components of Interventions. PLoS One. 2015;10(6):e0128581.

3. Hugtenburg JG, Timmers L, Elders PJ, Vervloet M, van Dijk L. Definitions, variants, and causes of nonadherence with medication: a challenge for tailored interventions. Patient Prefer Adherence. 2013;7:675–82.

4. Langendoen-Gort M, Al-Jabr H, Hugtenburg JG, Rutters F, de Wit M, Bhattacharya D, et al. A personalised intervention programme aimed at improving adherence to oral antidiabetic and/or antihypertensive medication in people with type 2 diabetes mellitus, the INTENSE study: study protocol for a randomised controlled trial. Trials. 2022;23(1):731.

5. Langendoen-Gort M, Rutters F, Huijts D, Elders PJ, Terwee CB, Hugtenburg JG. Validation of an Announced Telephone Pill Count Compared to a Home-Visit Pill Count in People With Type 2 Diabetes or Cardiovascular Disease. Clin Pharmacol Drug Dev. 2023;12(1):85–93.

6. Lam WY, Fresco P. Medication Adherence Measures: An Overview. Biomed Res Int. 2015;2015:217047.

7. Chan AHY, Horne R, Hankins M, Chisari C. The Medication Adherence Report Scale: A measurement tool for eliciting patients’ reports of nonadherence. Br J Clin Pharmacol. 2020;86(7):1281–8.

8. van der Laan DM, Elders PJM, Boons C, Nijpels G, van Dijk L, Hugtenburg JG. Effectiveness of a Patient-Tailored, Pharmacist-Led Intervention Program to Enhance Adherence to Antihypertensive Medication: The CATI Study. Front Pharmacol. 2018;9:1057.

9. van der Laan DM, Elders PJ, Boons CC, Bosmans JE, Nijpels G, Hugtenburg JG. The (cost-)effectiveness of a patient-tailored intervention programme to enhance adherence to antihypertensive medication in community pharmacies: study protocol of a randomised controlled trial. Trials. 2017;18(1):29.

10. van der Laan DM, Langendoen-Gort M, Nijpels G, Boons C, Elders PJM, Hugtenburg JG. Implementation fidelity of an intervention programme to enhance adherence to antihypertensive medication in Dutch community pharmacies. Int J Clin Pharm. 2019;41(4):1031–46.

11. Belete AM, Gemeda BN, Akalu TY, Aynalem YA, Shiferaw WS. What is the effect of mobile phone text message reminders on medication adherence among adult type 2 diabetes mellitus patients: a systematic review and meta-analysis of randomized controlled trials. BMC Endocr Disord. 2023;23(1):18.

12. Langendoen-Gort M, Bhattacharya D, Rutters F, Al-Jabr H, de Wit M, Hugtenburg JG, et al. Process evaluation of the INTENSE study aimed at improving adherence to oral diabetes or hypertension medication in people with type 2 diabetes mellitus. 2026. To be submitted.

